# Nutritional Status and Lifestyle of Children in Orphanages and Middle-Class Families in Khulna City, Bangladesh: A Cross-Sectional Study

**DOI:** 10.1101/2024.07.10.24310074

**Authors:** Tohora Safa, Mir Fhamid Ahsan Alvi, Maliha Mahazabin

## Abstract

The lack of biological parents has a significant effect on a child’s growth and overall welfare. This study examines the circumstances of children living in orphanages where one or both parents have passed away, as well as children from middle-class households. The main focus is on their lifestyle, nutritional status, and well-being. We performed a chi-square test and descriptive analysis to establish a correlation. Our research revealed that orphaned children in the orphanage tend to mostly consume carbohydrate-rich foods in their diet. Inadequate access to nourishing food can impede their capacity to sustain optimal physical and mental well-being. Furthermore, the absence of recreational amenities in the orphanage may impede their mental well-being, as they already experience psychological challenges due to the absence of parental affection. Children from middle-class households are often exposed to a significant amount of unhealthy fast food and bakery items. This practice should be discontinued in order to promote a healthier society.

## 1. Introduction

Approximately 140 million children worldwide were identified as orphans in 2015, according to UNICEF, which defines an orphan as a child under the age of 18 who has lost either of his or her parents due to a cause of death **[1]**. On the other hand, 2019 UNICEF research estimates that 24 million infants either live in alternative care or do not get any care from elders, and 130 million children worldwide are in danger of losing parental care **[2]**. There are approximately seven billion inhabitants on the earth, among them two and a half billion are children, and between 143 and 210 million are orphans. The continents with the highest rates of orphans include Asia, Africa, Latin America, and the Middle East **[1–3]**. Bangladesh has 4.8 million orphans. Both of their parents or just one of them passed away. Being vulnerable themselves, single parents are typically unable to maintain custody of their kids, particularly if they choose to remarry. Besides, Bangladesh faces the challenges of poverty, overcrowding, and natural catastrophes **[4]**.

According to estimates, there would be over 1.5 million motherless orphans in African countries, a country where HIV prevalence can reach 25%. Although it is customary in sub-Saharan Africa for extended households to care for orphans instead of placing them in orphanages, anecdotal data suggests that the growing number of HIV/AIDS orphans is straining community assets that are accessible. Eating a healthy diet is essential for optimizing brain performance and improving learning **[5]**. A piece of evidence depicts that childhood malnutrition, typified by stunted development, is linked with long-term inabilities in cognitive and academic performance, where social and psychological variables are absent. Moreover, there is risk factor which is called malnutrition that is increasing for children who are lagging by their parents **[5]**. One of the primary public health issues facing emerging nations is undernourishment in children under five [6]. Under nutrition, which includes stunting, wasting, underweight, and micronutrient deficiencies, is caused by insufficient consumption of both macro and micronutrients. Almost 149 million children under the age of five were stunted in 2018, and malnutrition was the primary cause of almost three-fourths of under-five mortality **[6]**.

There are between 8 and 10 million newborns and kids living in orphanages worldwide **[7]**. A previous study illustrates that dietary deficiencies are a common occurrence for children residing in orphanages worldwide. Asia and Africa are rife with orphan problems. Socioeconomically impoverished Asian nations have long practiced the practice of putting underprivileged children in orphanages **[7,8]**. Throughout the world, there are several “Resource-Constrained Communities,” or RCCs. A similarity across populations with limited resources, such as slum dwellers, beggars, orphans, etc., is that their numbers are notably greater in developing or third-world nations **[3]**. For instance, in Bangladesh, a developing nation that is expanding, there are around 7, 000, 00 beggars and 4, 000, 00 orphans. There are over 1,000,000 beggars in Dhaka, the capital of the country, and about 25 orphanages dispersed across the city. A large population of orphans reside in the orphanages **[3]**Young people comprise around 45% of the population of Bangladesh **[7]**. Bangladeshi children, like those in other least-developed countries, deal with several issues that might be detrimental to their psychological and physical well-being, which include malnutrition, filthy living conditions, hardship, contagious illnesses, lack of education, and broken houses **[7]**. Malnutrition and infectious infections plague children living in orphanages. A decrease in nutrition compromises the immune system, which in turn causes frequent and severe illnesses that further erode the intake of nutrients and perhaps endanger the life of the kid **[7,8]**. It is notable that among the children residing in Bangladeshi orphanages, malnourishment, physical or psychological abuse, food insecurity, and a dearth of parental supervision and protection are the most prevalent conditions **[7]**.

Ninety-five percent of the projected 132 million single and double orphans reside in low- and middle-income countries (LMICs). On top of that, care is required for hundreds of millions of street kids, whose numbers are rising throughout several nations **[9]**. The word “institutions,” which is used generally in policy and program planning papers to describe group homes where children from many families live with biologically unrelated caretakers, is projected to be the home of more than two million children **[9]**. These group homes are diverse; they range from modest houses with long-term live-in caretakers to enormous establishments with several family-like units to huge establishments with caregivers working in shifts and without specific child-related responsibilities **[9]**. Orphans and middle-class family infants lead very different lifestyles as their family support, financial status, and availability of resources are different. To begin with, Middle-class families typically provide security with better access to medical care, schooling, and recreational activities **[10]**. On the contrary, volatility, limited access to resources, and mental health problems are frequent challenges faced by orphans, which harm their overall development and wellbeing. Secondly, Middle-class families usually raise stable, easily accessible children who enjoy easy access to basic needs. Although their earnings are often secure, numerous households in this category can afford greater comfort, more nutritious food, and further opportunities Moreover, improved psychological and physical health results are often attained when health care is consistently available **[10–12]**. Besides, these children have a vast development in their lifestyles as they have strong social networks as well as regular participation in extracurricular activities. Orphans, on the other hand, usually have rather different lifestyles. Many orphans face a lot of problems including inadequate nutrition, poor healthcare, and limited educational opportunities as some of them live in institutions and some of them with their families **[12]**. If their parents fail to provide them with the care and support they need, they could have emotional and psychological issues that hinder their growth. Such children are more likely to experience malnutrition, maltreatment, and poor school performance, all of which affect their overall development **[10,11]**.

There are in total of 169.4 million people in Bangladesh as it shows from research of 202. About 40 percent of the total population is composed of over 64 million infants. Fifty percent of Bangladesh’s 64 million youngsters live in poverty and impoverishment. The orphans’ situation is far worse **[3,4]**. While some government and NGO charities offer minimal housing and assistance, their finances are severely constrained, and they lack a robust support network for extracurricular activities, education, healthcare, and counseling **[5,12]**. There are several changes in the lives of an orphan and a child from a middle-class family in Bangladesh. Bangladeshi middle-class families can afford some utilities for their children whereas an orphan can’t get those utilities including caring environment, security, healthcare & nutrition, and so on. It is more usual for these children to live in safe families, participate in extracurricular activities, and get the emotional and material support they need to flourish. The consistency and predictability this environment offers are essential to their physical and mental growth. However, some challenges commonly obstruct the development and welfare of Bangladeshi orphans. Afterward, the orphans of Bangladesh face many challenges which commonly obstruct their development. Most of the orphans live in temporary residences or with extended family members who may not have enough funds to provide them with the care, security, healthcare & nutrition they need. They frequently experience high rates of malnutrition, limited access to education, and terrible living conditions. Most of the time of their lifestyle, these orphans suffer psychologically as a consequence of their chaotic living conditions and the unfortunate passing of their parents. These children face some adjusting problems to society and achieving scholastic objectives due to a lack of support and proper guidance from their parents. The variations in the lifestyle between the orphans and middle-class family children depict the signific of the support from their family, malnutrition problems, and socioeconomic stability for children’s healthy growth. Our study observed the socioeconomic and nutritional status of middle-class family children and orphan children. Our research aims to explore orphan children’s socioeconomic and nutritional status and compare their situation with middle-class family children.

## 2. Materials and methods

The research was carried out in several areas of Khulna city. This study examined the nutritional status of orphaned children and school-going children from middle-class families. The age range of the participants in this study was from 6 to 14 years old. The primary data collection included a systematic random sampling technique. The total sample size is 250. Among them 125 are orphan children and 125 are school going children from middle class families. Prior to conducting the actual interview, the study involved explaining the aims and objectives to the respondents in person. This was done to ensure that they understood the purpose of the study and felt comfortable speaking openly. Subsequently, the inquiries were presented straightforwardly, accompanied by clarifications if deemed essential. Time was conserved by consistently avoiding irrelevant inquiries. To mitigate the influence of respondents’ recall bias, repeated visits were made and questions were presented in a systematic order, facilitating the recall of relevant information.

The anthropometric data (weight, Height and MUAC (mid-upper arm circumference)) of the Children were taking individually; weight was recorded Kilogram and height in centimeter.

Age group 6-9
| MUAC Range | Nutritional Status |
| --- | --- |
| <13.5-15.5 | Severe |
| 15.5-18.5 | Moderate |
| 18.5-23 | Normal |

Age group 10-14
| MUAC Range | Nutritional Status |
| --- | --- |
| <16-18.5 | Severe |
| 18.5-23 | Moderate |
| 23-32 | Normal |
| >32 | Overweight |

We completed pre-tabulation tasks before the final tabulation to ensure their usefulness for analysis and to obtain reasonable information. We carefully edited the collected raw data to detect errors and omissions, and to eliminate irrelevant information. In this study, descriptive methods were primarily used for data analysis. Data were analyzed through univariate methods, descriptive statistics, and Pearson’s Chi-square test using IBM SPSS software.

## 3. Results

In this study Table 1 shows Socio-demographic and lifestyle-related characteristics of orphan children. About 50.4 % of orphan children said that they come to the orphanage when their age is within 3-6 years. 36 % of orphan children said that they come to the orphanage when their age is within 7-10 years. 13.6 % of orphan children said that they come to the orphanage when their age is between 11-14 years. The survey revealed that of all the orphan children 20 (25%) said that 3-6 lives in a room. 39 (31.2%) children said that 7-10 lives in a room and 31 (24.8%) %) children said that 11-14 lives in a room. On the other hand 30 (24%) %) of children said that 15≥ live in a room. Approximately 77 % of orphan children said that they get sufficient space in their room and 22.4% of orphan children said that they cannot get sufficient space in their room. Most of the orphan children 50 (40%) said that they cannot get good treatment facilities during the sick period. But 28 (22.4%) said that they could get treatment facilities during the sick period and sometimes 47(37.6%) said that they could get treatment facilities during the sick period. Furthermore, the survey revealed that most of the orphan children 79(63.2%) said that their entertainment tools were sufficient and 46(36.8%) said that their entertainment tools were not sufficient. On the other hand, 106(87.2%) middle-class families’ children said their school has sufficient entertainment tools and 19 (15.2%)said that their entertainment tools are not sufficient. We found that 44(35.2%) orphan children said the authority did not take any steps to go outside for mental recreation and 81(64.8%) said sometimes the authority took to go outside for mental recreation. Here, 76(60.8%) middle-class families’ children said their school took to go outside for mental recreation and 49(39.2%) sometimes the authority took to go outside for mental recreation. The survey revealed that 57(45.6%) said socially they can communicate with other children. 33(26.4%) said socially they can communicate with other children and 35 (28%) said sometimes they can communicate with other children. We found that 11 (8.8%) orphan children said that they can get opportunities to play facilities outside of the orphanage and 74(59.2 %) children said that they cannot get play facilities outside of the orphanage.

**Table 1.** Socio-demographic and lifestyle-related characteristics of orphan children.

| Factors | Number of Respondents | Percent |
| --- | --- | --- |
| **Admitted Age in the Orphanage** |  |  |
| 3-6 | 63 | 50.4 |
| 7-10 | 45 | 36 |
| 11-14 | 17 | 13.6 |
| **Number of Orphan live in a room** |  |  |
| 3-6 | 25 | 20 |
| 7-10 | 39 | 31.2 |
| 11-14 | 31 | 24.8 |
| 15≥ | 30 | 24 |
| **Sufficient living space** |  |  |
| Yes | 97 | 77.6 |
| No | 28 | 22.4 |
| **Have treatment facilities** |  |  |
| Yes | 28 | 22.4 |
| No | 50 | 37.6 |
| **Special food and nursing facilities for sick orphan children** |  |  |
| Yes | 26 | 21 |
| No | 99 | 79 |
| **Description of the respondent by doing when they missing the parental love** |  |  |
| Crying | 28 | 22.4 |
| Keep Silent | 65 | 52.0 |
| Nothing to do | 32 | 25.6 |
| **Social Communication among orphan with other children Percent** |  |  |
| Yes | 57 | 45.6 |
| No | 33 | 26.4 |
| Sometimes | 35 | 28 |
| **Outgoing Playing Facilities of Orphan Children** |  |  |
| Yes | 11 | 8.8 |
| No | 74 | 59.2 |
| Sometimes | 40 | 32 |
| **Rehabilitated after release from the orphanage** |  |  |
| Hand over Guardian | 82 | 65.6 |
| Higher study | 9 | 7.2 |
| Others | 34 | 27.2 |

Table 2 shows that among the respondents, 65.6 % of orphan children and 59.2 % of middle-class families’ children don’t share food with their friends. On the other hand, 14.4 % of orphan children and 9.6 % of middle-class families’ children like to share food with their friends and 20 % of orphan children and 31.2 % of middle-class families’ children sometimes share food with their friends. The study indicates that almost all orphanage children 50 % of them do not know about nutritional requirements. But the rest of the middleclass families’ children 6 % have little idea about nutrition. Most of the orphan children 89.6 % intake carbohydrates so they get nutrients mainly carbohydrates and only 8% intake protein and 2.4 % intake others. On the other hand, middle-class families’ children have 24 % little amount of carbohydrate intake and most middle-class families’ children 60% intake of protein and 16 % intake of others. We found that 22.4% of orphans and 13.6% of middle-class families of school children of our studied respondents have a BMI of less than 16 and MUAC of less than 13 a result that suggests a high risk. 53.6 % of orphans and 32 % of middle-class families’ school children of our studied respondents are moderately malnourished. On the other hand, only 24 % of orphans and 52.8 % of middle-class families’ school children are well nourished.

**Table 2.** Lifestyle, knowledge and nutrition-related characteristics Orphan children And Children of Middle Class Families.

| Factors |  |  |
| --- | --- | --- |
| Children Share Food | Percent |  |
|  | Orphan Children | Children of Middle Class Families |
| Yes | 14.4 | 9.6 |
| No | 65 | 59.2 |
| Sometimes | 20 | 31.2 |
| Children had Knowledge about Nutrition | Percent |  |
|  | Orphan Children | Children of Middle Class Families |
| Yes | 0 | 6 |
| No | 50 | 44 |
| Children had sufficient entertainment tools or instrument | Percent |  |
|  | Orphan Children | Children of Middle Class Families |
| Yes | 63.2 | 87.2 |
| No | 36.8 | 15.2 |
| Authority takes any steps to go outside for mental recreation | Percent |  |
|  | Orphan Children | Children of Middle Class Families |
| Yes | 0 | 60.8 |
| No | 35.2 | 0 |
| Sometimes | 64.8 | 39.8 |
| Description of the Behavior Teacher or Authority | Percent |  |
|  | Orphan Children | Children of Middle Class Families |
| Well | 25.6 | 54.4 |
| Excellent | 0 | 20 |
| Authorial | 74.4 | 25.6 |

| Sources of Nutrient Intake | Percent |  |
| --- | --- | --- |
|  | Orphan Children | Children of Middle Class Families |
| Carbohydrate | 89.6 | 24 |
| Protein | 8 | 60 |
| Others | 2.4 | 16 |
| Amount of Calorie Intake per Day | Percent |  |
|  | Orphan Children | Children of Middle Class Families |
| 951-1450 cal | 22.4 | 13.6 |
| 1451-1950 cal | 53.6 | 35.2 |
| 1951-2450 cal | 24 | 51.2 |
| Assessment of Nutritional Status by Using MUAC | Percent |  |
|  | Orphan Children | Children of Middle Class Families |
| Severe | 22.4 | 13.6 |
| Moderate | 53.6 | 32 |
| Normal | 24 | 52.8 |
| Overweight | 0 | 1.6 |

From Figure 2, it can be seen that among the respondents about 20.4 % of respondents belong between the age 6-8 years, 42.8 % belong between the age 9-11 years 36.8% belong between the age 12-24 years. Moreover from Figure 3, it can be seen that Most of the orphan children’s height is between 161-170 cm and most of the middle-class families children’s height is between 131-140 cm. Most of the orphan children and middle class families’ children weight is between 36-45 kg.

**Figure 1.**
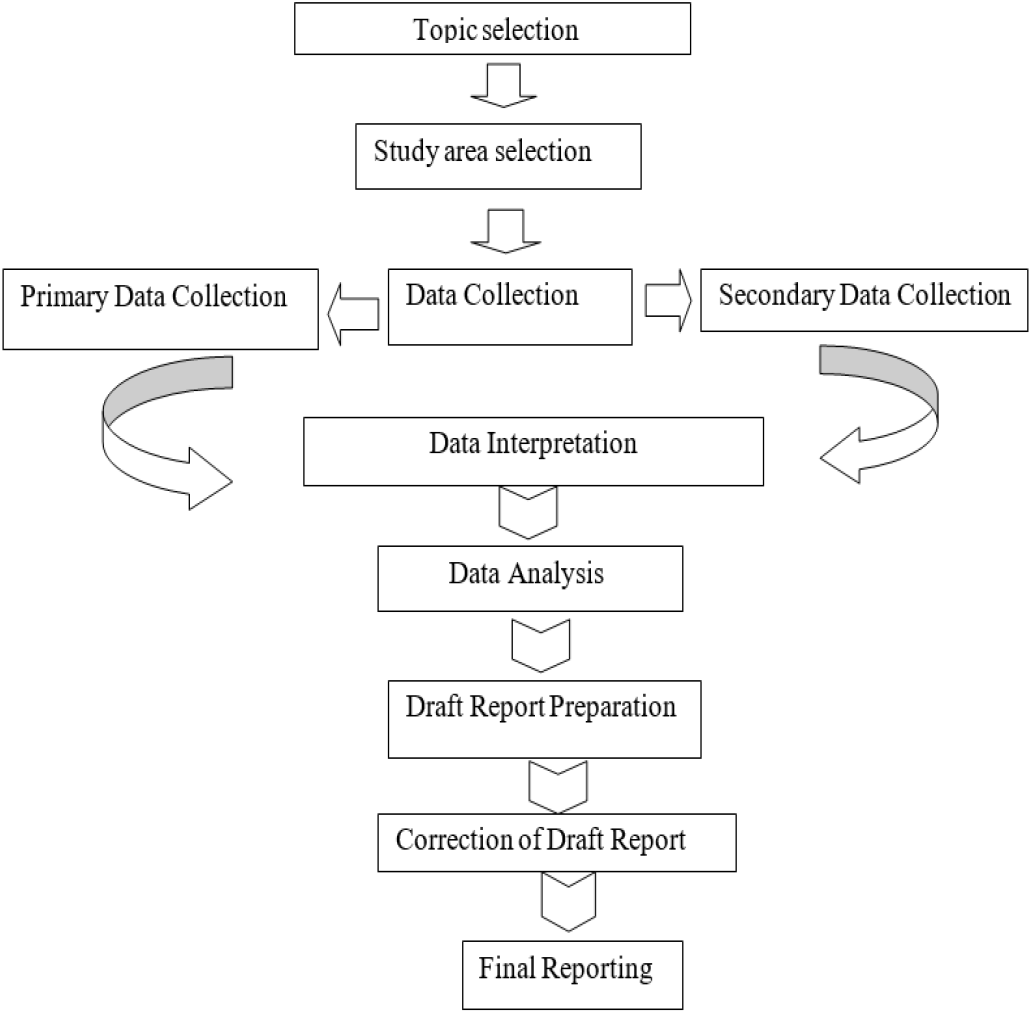
Flow Chart of the phases of Research Activities

**Figure 2.**
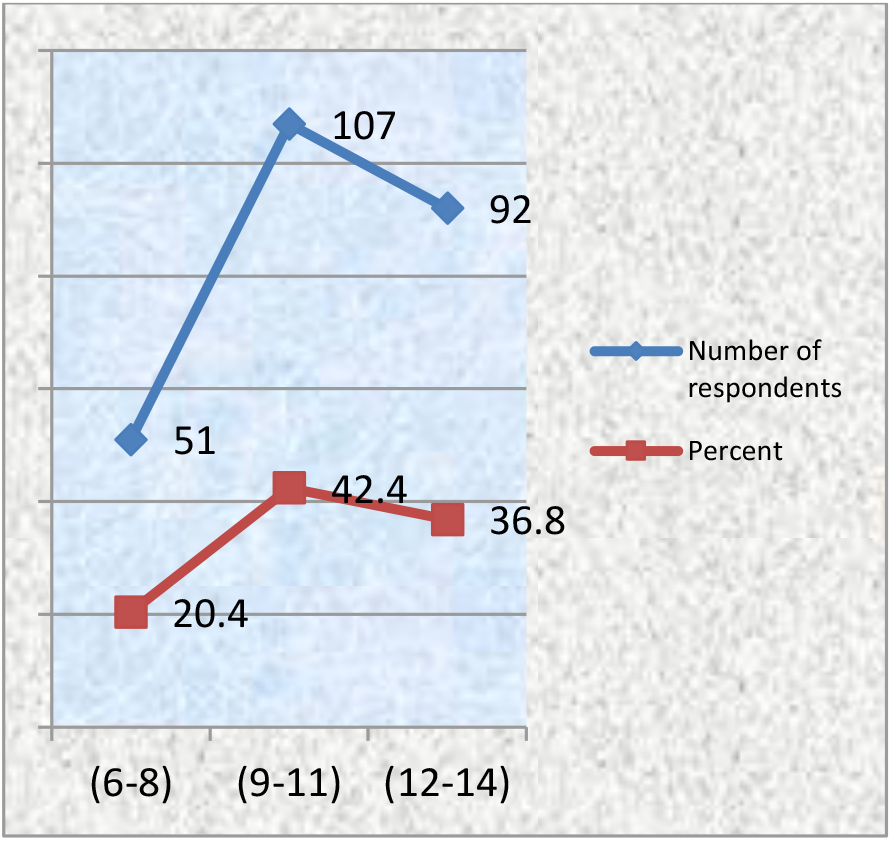
Distribution of Respondents by Age

**Figure 3.**
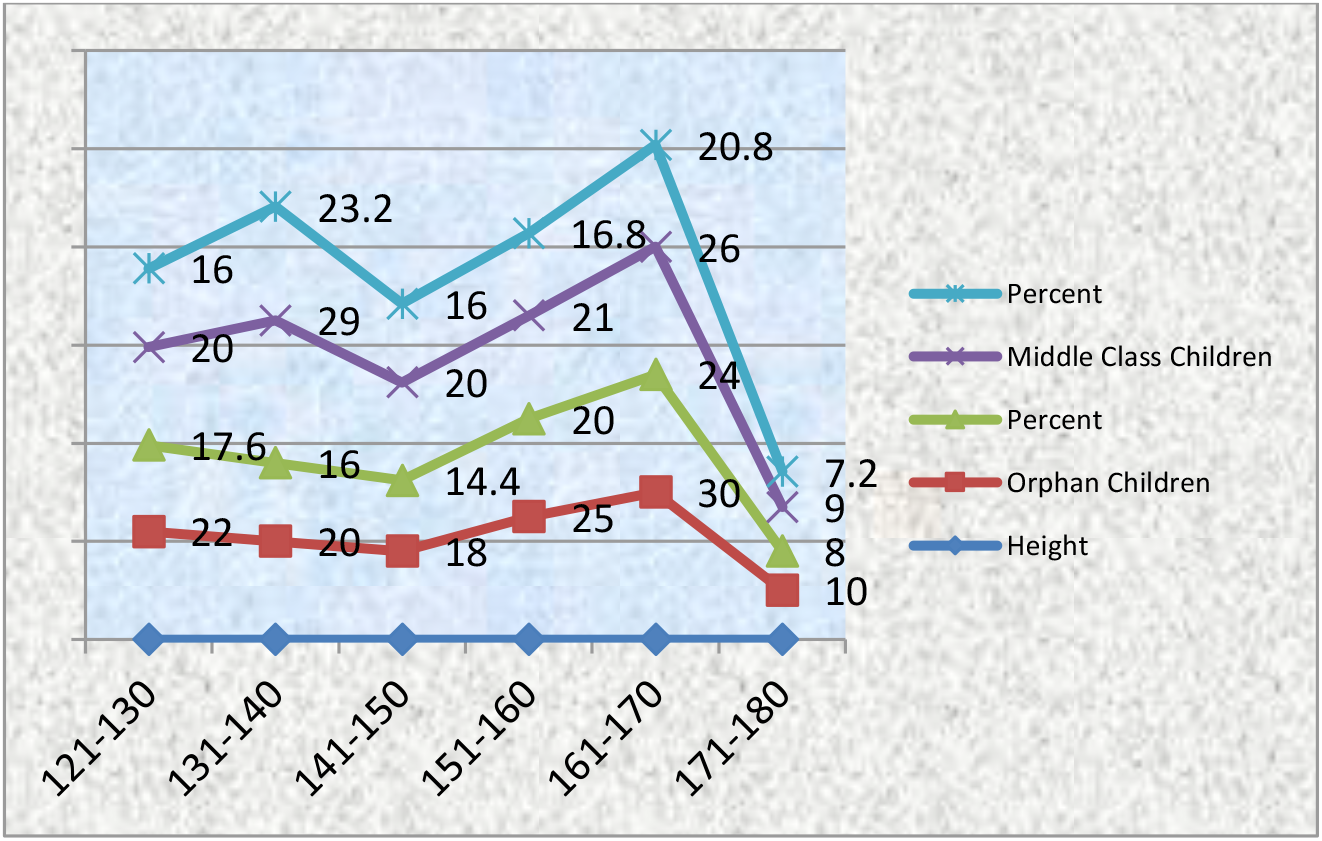
Distribution of Respondents by Height

**Figure 4.**
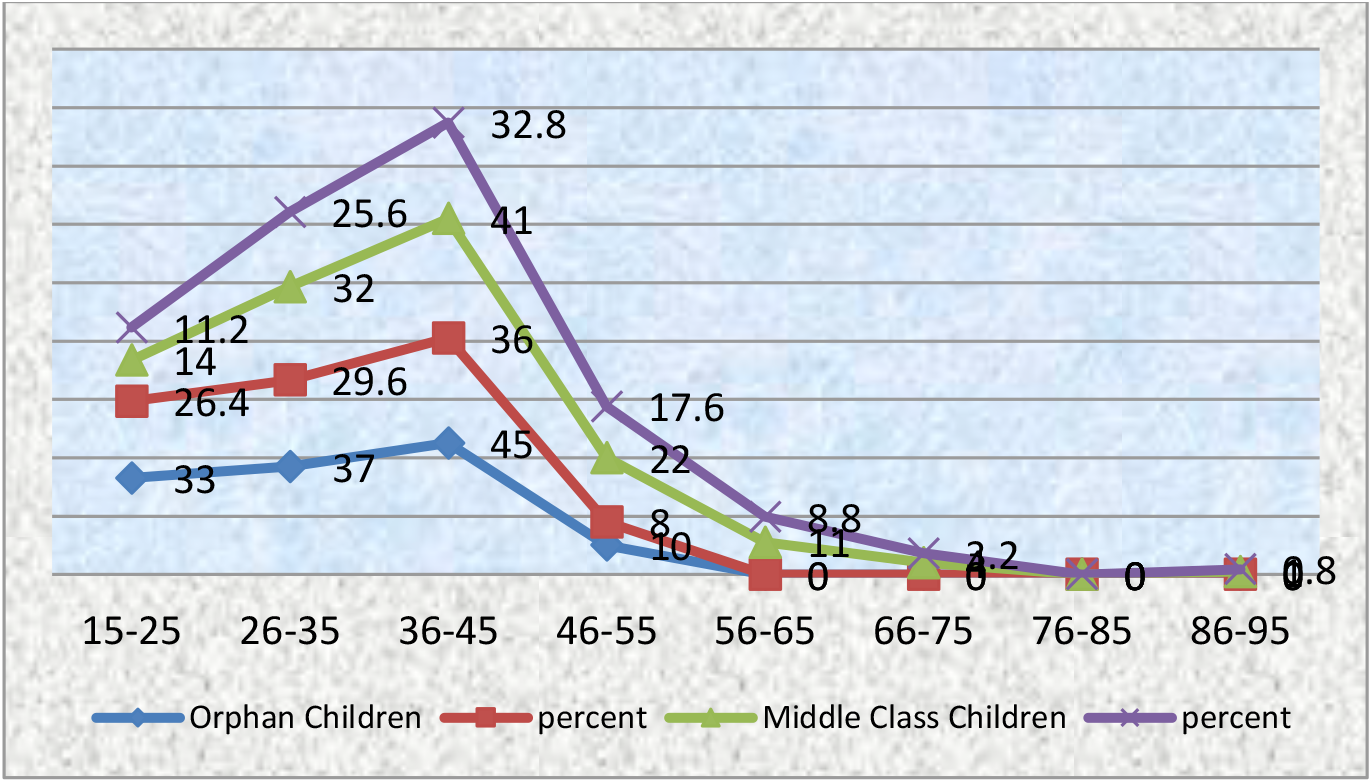
Distribution of Respondents by Weight

## Correlation of different variables

### Nutritional knowledge and their Daily Calorie Intake by Orphan Children

**H**_**a**_: There is a relation between the nutritional knowledge of orphan children and their daily calorie intake.

**H**_**o**_: There is no relation between the nutritional knowledge of orphan children and their daily calorie intake.

Table 3 indicates that the calculated value of Pearson’s *X*^2^ is 2.500 at 3 degree of freedom with 0.01 percent level of significance. The null hypothesis is rejected, since p < 0.05 (in fact p < 0.001). Here, p= the value of Exact. Sig.

**Table 3.** Nutritional knowledge of orphan Children and their Daily Calorie Intake by Orphan Children.

| Nutritional Knowledge of Orphan Children | Intake of Daily Calorie |  |  | Total |
| --- | --- | --- | --- | --- |
|  | 951-1450 | 1451-1950 | 1951-2450 |  |
| No | 28 | 67 | 30 | 125 |
|  | 22.4 | 53.6 | 24.0 | 100.0 |
| Total | 28 | 67 | 30 | 125 |
|  | 22.4 | 53.6 | 24.0 | 100.0 |
| Pearson's $X^2 = 2.500$ | | Degree of Freedom=3 | Level of Significance=.01% | P < .001 |

### Educational Qualification of the Respondents Parent and Nutritional Status of the Children of Middle Class Families

**H**_**a**_: There is a relation between educational qualification of respondents’ parent and nutritional status the children of middle class families.

**H**_**o**_: There is no relation between educational qualification of respondents’ parent and nutritional status the children of middle class families.

Table 4 indicates that the calculated value of Pearson’s*X*^*2*^ is 2.503 at 8 degree of freedom with 0.01 percent level of significance. The null hypothesis is rejected, since p < 0.05 (in fact p < 0.001). Here, p= the value of Exact.

**Table 4.** Educational Qualification of the Respondents Parent and Nutritional Status of the Children of Middle Class Families.

| Educational Qualification of the Respondents Parent | Nutritional Status |  |  |  | Total |
| --- | --- | --- | --- | --- | --- |
|  | Severe | Moderate | Normal | Overweight |  |
| Educated | 11 | 25 | 41 | 1 | 78 |
|  | 14.10 | 32.05 | 52.5 | 1.2 | 62.4 |
| Higher Educated | 6 | 15 | 25 | 1 | 47 |
|  | 12.7 | 31.9 | 53.19 | 2.12 | 37.6 |
| Total | 17 | 40 | 66 | 2 | 125 |
|  | 13.6 | 32.0 | 52.8 | 1.6 | 100.0 |
| Pearson's $X^2 = 2.503$ | Degree of Freedom=8 | | Level of Significance=.01% | | P < .001 |

### Nutritional knowledge and their Daily Calorie Intake by Children of Middle Class Families

**H**_**a**_: There is a relation between nutritional knowledge about middle class Children and their daily calorie intake.

**H**_**o**_: There is no relation between nutritional knowledge about middle class Children and their daily calorie intake.

Table 5 indicates that the calculated value of Pearson’s *X*^*2*^ is 2.521 at 6 degree of freedom with 0.01 percent level of significance. The null hypothesis is rejected, since p < 0.05 (in fact p < 0.001). Here, p= the value of Exact. Sig.

**Table 5.** Nutritional knowledge and Calorie intake by Children of Middle Class Families.

| Nutritional Knowledge of Middle Class Children | Intake of Daily Calorie |  |  | Total |
| --- | --- | --- | --- | --- |
|  | 951-1450 | 1451-1950 | 1951-2450 |  |
| Yes | 3 | 6 | 6 | 15 |
|  | 20.0 | 40.0 | 40.0 | 12.0 |
| No | 14 | 38 | 58 | 110 |
|  | 12.72 | 34.5 | 52.7 | 88.0 |
| Total | 17 | 44 | 64 | 125 |
|  | 13.6 | 35.2 | 51.2 | 100.0 |
| Pearson's $X^2 = 2.521$ | Degree of Freedom=6 | Level of Significance=.01% | | P < .001 |

## 4. Discussion

Our study observed the socioeconomic and nutritional status of middle-class family children and orphan children. Our research aims to explore orphan children’s socioeconomic and nutritional status and compare their situation with middle-class family children. Our study found that 22.4% of orphans and 13.6% of middle-class families’ schoolchildren have a BMI of less than 16 and a mid-upper arm circumference of less than 13, indicating a high risk. Approximately 32% of school-age children from middle-class families and 53.6% of orphans in our study had moderate malnutrition. However, only 24% of orphans and 52.8% of middle-class families’ school-age children were well nourished. Previous studies in Bangladesh’s Dhaka city revealed that mild, moderate, and severe malnutrition affected 43.1%, 16.8%, and 0.4% of the children at the orphanage, respectively **[7]**. Moreover, other institutionalized school-age orphans in Harari Regional State in eastern Ethiopia reported that 15.8%, 10.9%, and 8.7% of the orphans had stunting, wasting, and underweight, respectively **[13]**. Another study conducted in Bangladesh revealed that 60.87% of orphans attending school had a normal weight, 21.74% were classified as overweight, 6.5% were classified as obese, and 10.87% were classified as underweight **[7]**. In our study, about

50.4 % of orphan children said that they came to the orphanage in their early childhood (3 to 6 years old), and 36 % of orphan children said that they come to the orphanage when their age is within 7-10 years. Only 13.6 % of orphan children said they came to the orphanage in their early adolescence (11 to 14 years old). Previous studies conducted in West Africa (Ghana) found that 4.9% of orphan children said that they came to the orphanage in their early childhood (0 to 5 years old), and 4.1 % of orphan children said that they come to the orphanage when their age is within 6-11 years. However, 22% of the orphan children came here at the age of 11 to 23 **[14]**. Moreover, a study was conducted in Tangail, Bangladesh with orphan 200 children, where around 33% of them were in their early adolescence (12 to 14 years old), and 12.5% of them were 15 to 17 years old **[15]**. Our study shown that, majority of respondents were girls (70.4 %), whereas only 29.6 % of the children were boys. Previous studies conducted in West Africa (Ghana) found that the majority of the children were male (52.4%) **[14]** and another study in Dhaka city in Bangladesh, found that more than half (56.0%) sample were boys **[7]**. We found that among middle-class family children, only 22.4% of children preferred the value of nutritious homemade food. Previous study also mentioned similar findings which reported that high levels of school going children eat healthy, following dietary rules, usually at homes **[16]**. Although a significant portion of the population, 43.2%, prefers bakery and fast food. Previous research has indicated that, although our students used to enjoy eating meals prepared at home, they no longer do. Other research has revealed that, in Bangladesh, children today prefer and desire to eat “better food,” which includes meat, fast food, sugary drinks, burgers, chips, and other fried and sweet foods available in the market. These foods are unhealthy and can contribute to obesity if consumed on a regular basis **[8]**. We found that approximately 40% of orphan children said all the demands fulfill the authority and the majority (73%) of orphan children said that they cannot get choice full food, which aligns with previous studies **[17,18]**.

We found that the majority of orphan children (89.6%) consume carbohydrates, so they obtain nutrients primarily from carbohydrate-based foods; only 8% consume protein-related foods. While middle-class families’ children consume a small amount of carbohydrates (24%), with the majority of them consuming proteins in their diets. In previous studies the average intake of energy, protein, carbohydrates, and fat was found to be 2270 kcal, 65 grams, 335 grams, and 73 grams, respectively. Protein, fat, and carbohydrates make up 59%, 12%, and 29% of total calories, respectively. The protein consumption was 65 grams, which is over 50% more than the 43.40 grams required amount. An urban nutrition study reported that when it came to calorie consumption, it was discovered that the group consumed roughly 20% more than what was needed and 42% more than what was reported **[7]**.In other studies in Kenya 45.2% fewer children received food from four or more food categories. These findings are consistent with research conducted in Pakistan that discovered 42.2% of school-age children ingested four or more food groups. Compared to children who ate from less than four food categories, those who ingested items from four or more food groups were at a lower risk of stunting and underweighting. Eating a diverse diet has been linked to improved health and an increase in energy intake. Due to their bulk and satiety value, starchy cereals can be fed in big portions to children **[19]**. The study participants, who were kids and adolescents, had repetitive meals that lacked animal-derived protein, and a variety of vegetables and fruits. The likelihood of iron sufficiency was elevated, which can lead to stunting, underweight, and adolescent overweight when carbohydrate intake is high. Inadequate calorie and iron consumption can result in stunted growth and underweight, respectively **[13,14,20]**.

Our research revealed that the majority of orphanages do not maintain adequate hygiene. However, the majority of schools only moderately maintain their hygienic conditions, while some maintain theirs properly. Eventually, previous study 81.5% of the 200 orphanage children in Tangail, Bangladesh, were found to regularly bathe, 89.5% to brush their teeth, 97.5% to wash their hands before eating, 72.5% to wash their clothes, 64.0% to clip their nails, and 97% to wear sandals **[15]**. The aforementioned indicators are indicative of improved personal hygiene. In our study, we found that majority (85%) of children do not have idea about the importance of washing hand before and after taking food .and the study shows that usually they are not habitually wash their hand. According to earlier research on hygiene and health-related behavior, 240 children (90.6%) and 93.6%, respectively, frequently washed their hands after using the restroom and before eating. Only 51.3% said they washed their hands with soap. Forty (15.1%) of the children’s caregivers said that they had recently fallen ill most probably due to unhygienic behavior **[13,17]**. We found, more than 30% of orphan children reported that, in the orphanage 7 to 10 children live in a room and only 7.6% orphan children said that they get sufficient space in their room. In the orphanage, 24.8% of children said that the same age group of orphan children live in their room. Approximately 44% of children reported that their room was properly ventilated aligning with previous study findings **[21]**. Most of the orphan children (40%) said that they could not get well treatment facilities during the sick period and all of the orphan children said that, they were taken to the hospital immediately at a serious moment. Only 21% of orphan children reported that they get special food and nursing facilities when they get sick in the orphanage, which aligns with previous studies **[13,22–24]**. We found that majority of children (60%) in the middle class family reported that the school authority takes steps to go outside for mental Recreation. However, no orphan children reported that the orphanage authority takes steps to go outside for their mental recreation, which coincides with prior studies **[25–27]**. We found that more than half (52%) of orphan children said they remain silent when they miss their parents’ love. Only 22% of orphaned children cry when they think about or miss their parents **[28–30]**. Children’s basic attachment requirements, as well as their specific emotional needs as orphans, must be addressed.

## 5. Conclusion

As a developing nation, Bangladesh is confronted with numerous socio-economic challenges across various sectors, including food security, nutrition, education, healthcare, and gender inequality. Malnutrition primarily affects developing countries. Malnutrition can result from inadequate dietary choices. Malnutrition can occur in children who are either undernourished or overnourished. The primary focus of national life should be the provision of adequate nourishment, education, healthcare, clothes, and shelter for children. Children are the prospective leaders of a nation. This study aims to evaluate and compare the general health condition, lifestyle, and nutritional status of orphan children and children from middle-class homes. Children from middle-class households have a preference for indulging in delicious snacks that are detrimental to their health. Consequently, their weight is unexpectedly increasing, with only a mere 6% being exempt from this trend. Children from middle-class backgrounds has less knowledge on nutrition. Approximately 60% of youngsters from middle-class families consume nutritious food. However, orphaned children lack knowledge on nutrition. What is noteworthy is that a majority of children from middle-class families have a preference for fatty foods. Children do not adhere to a regular meal schedule. Orphaned children face detrimental effects including inadequate sanitation, substandard hygiene practices, insufficient food intake, heightened vulnerability to illness, and a lack of proper nutrition. Efforts should be made to promote their cognitive development and ensure that their fundamental needs are met. In order to address the issue related to the neglected constructive approach towards all sectors of society, it is imperative for the appropriate advancement of the future of orphans that every sector of society actively participates.

## Recommendations

### Orphan Children

- The number of orphanage should be increased.
- Sufficient balanced diet for the children
- They must be provided good medical care.
- They must be provided with the equal social facility as mass people.
- They must be facilitated with education, specially private tuition.
- For the development of orphans and orphanages provision of special fund is essential.

### Children of Middle class families

- Middle class families’ children should avoid fast food.
- Every school should have playing ground for the children
- Every school must be maintain proper sanitation.
- Eat their meals proper time.
- Pack home prepared food.

## Data Availability

Data will be available upon reasonable request.

## Author Declarations

This work did not receive any fund. The authors declared no competing interest. All relevant ethical guidelines have been followed, the study received approval Ethical Clearance Committee review board to ensure compliance with ethical principles and safeguard the rights of participants. Before conducting the interviews, all participants were provided with information about the study and were also informed about the confidentiality and anonymity of their responses.

## Notes

### Competing Interest Statement

The authors have declared no competing interest.

### Funding Statement

We did not receive any funding for the research.

### Author Declarations

All relevant ethical guidelines have been followed, the study received approval from the Khulna University Ethical Clearance Committee review board to ensure compliance with ethical principles and safeguard the rights of participants. Prior to the survey, the following consent was obtained from the guardian. “I, the participant’s father, mother, or duly appointed guardian (orphanage), possess full authority, and I voluntarily consent for the child to participate in this study (if they assent to their participation) and understand that both I and the children have the right to refuse to answer questions.”

